# Insights into Parental Behavior: Self-Medication Patterns with antibiotics for Child Health

**DOI:** 10.1101/2024.03.20.24304568

**Authors:** Shishir Kumar, Shivani Agrawal, Tajwar Yasmin, Setu Sinha

## Abstract

**Objectives:** The alarming trend of self-medication with antibiotics in children, exacerbated by factors such as easy access to medications and insufficient awareness of the consequences, presents a critical health concern. This study aims to understand the behavior of parents on self medicating their children with antibiotics.

**Methods:** A cross-sectional study was done in the community under UHTC of IGIMS, Patna among 173 parents having children <12 years of age for a period of 12 months. Participants were selected through simple random sampling and were interviewed using a questionnaire developed by the authors. Data were compared using logistic regression and presented with odd ratios and confidence intervals.

**Result:** Prevalence of self medication was 31.8% in this study. Higher prevalence was seen among mothers, parents between 30-39 years, graduates (p = 0.001), having family income 20000-40000, housewives and those who do not have Ayushman card. Mothers, parents ≥40 years, having Secondary/ Higher secondary education, not having Ayushman card and those having child’s age of 7-9 years normally stop giving antibiotics when their child start feeling better. Whereas postgraduates parents (p = 0.000) and participants having family member in medical field were seen keeping antibiotic stock at home for later use. 39.9% parents could identify antibiotics correctly. Majority of the parents wait for 1-2 days before starting antibiotic. Most common reason for self medication was minor illness followed by previous experience with similar symptoms.

**Conclusion:** Lack of essential knowledge about medicines among parents is a serious matter particularly when children are concerned. Interventions targeted at improving awareness about antibiotic misuse, resistance and adverse effects amongst parents involved in self-medication need to be implemented on large scale.

## Introduction

Practicing self-medication with antibiotics (SMA) is on a rising trend in both developed and developing countries in recent years, being one of the most dangerous and inappropriate antibiotic use behaviors.^1^ Self-medication is defined as “the use of drugs to treat self-diagnosed disorders or symptoms or intermittent or continued use of a prescribed drug for chronic or recurrent disease or symptoms without a valid prescription.^2^ Inappropriate use of antibiotics is the key factor to the development of resistance which endangers their therapeutic effectiveness, increases treatment failures and leads to more severe illness episodes with higher costs and mortality rates.^3^

A meta-analysis with 57 studies indicated that the prevalence of SMA among children was 24% worldwide, with higher prevalence in the Middle East at 34%, Africa 22%, Asia 20%, and South America at 17%, while the lower prevalence in Europe at 8%.^4^ The prescribing patterns of antibiotics are not well controlled in many countries especially the developing ones which emphasizes the need to investigate and tackle such unhealthy practices.^5^ As there are fewer studies on self-medication with antibiotics in children in India, we aim to assess the behavior of parents on self-medicating children with antibiotics.

## Material and Methods

### Setting and design

A cross-sectional study was done for a period of 12 months in the community under UHTC of IGIMS, Patna among 173 parents having children below 12 years of age residing in the study population area selected through simple random sampling.

### Data collection procedure

Before initiating the enrolment process, the investigators informed the participants about the purpose of conducting the study. Strict confidentiality was ensured to all the participants and informed consent was taken individually prior to data collection. Data was collected using semi-structured, pre-tested, pre-coded questionnaire. House-to-house visit was done and participants were randomly selected. The participants were interviewed face to face and the responses were recorded in the master data sheet. The questionnaire was filled in two parts, first part contains socio-demographic characteristics and second part consists of behavior regarding self-medication with antibiotics in children aged 0-12 years. Participants who gave consent for participation, had children <12 years of age and parents having more than 1 child of age <12 years were enrolled only once in the study. Non-cooperative, non-consenting participants and those not residing in the study area for >6 months were excluded from the study. The study started after receiving approval (Letter No./807/IEC/IGIMS/2022) from the Institutional Human Ethics Committee and the Dean of Research at the Indira Gandhi Institute of Medical Sciences, Patna.

### Statistical analysis used

First, we coded and entered the data into a Microsoft Excel. We then used a computer software program PSPP to conduct our analysis. To understand the socio-demographic characteristics of our participants, we employed frequencies and percentages. This helped us to get a clear picture of the distribution of different demographic factors among our sample.

In addition, we employed logistic regression coefficient analysis. This allowed us to estimate odds ratios for independent variables and to identify predictors related to various aspects of behavior concerning self-medicating children with antibiotics. We considered a p-value of less than 0.05 to be statistically significant, indicating a strong association between variables.

## Patient and public involvement

The participants were involved after receiving approval from ethics committee. The purpose of the study was communicated to all the participants. However, participants were not directly or indirectly involved in the design of the study.

## Result

A total of 173 parents were included in the study. Prevalence of self medication with antibiotics in children was found to be 31.8% out of which 24.9% mothers and 6.9% fathers admitted that they give antibiotics to their children without prescription. Higher prevalence was found among parents between 30-39 years age group (15.6%), graduates (20.2%), having family income between 20000-40000 (19.7%) and those who were housewives (17.9%). Prevalence was significantly high among those who do not have Ayushman card (31.2%) and those having children of 4-6 years age.

On enquiring about the behavior of parents towards self medicating their children, mothers, parents ≥40 years age, having Secondary/ Higher secondary education, not having Ayushman card and those having child’s age of 7-9 years (OR: 1.034, 95% CI: 0.308 – 3.467; p – 0.957) normally stop giving antibiotics when their child start feeling better. Whereas parents of <30 years, doing service, postgraduates (OR: 12.550, 95% CI: 3.192 – 49.338; p – 0.000), having Ayushman card, and having family member working in medical field were seen keeping antibiotic stock at home for later use.

Table 3 shows multiple responses on whether parents could identify antibiotic correctly, how long they wait before starting antibiotic when their children get sick and reasons for self-medication. Of the total 173 parents only 69 (39.9%) parents could identify antibiotics correctly. Majority of the parents (85%) wait for 1-2 days before starting antibiotic. Most common reason for self medication was minor illness (25.4%) followed by previous experience with similar symptoms (22%). Only 01 parent stated high cost of medical consultation as the reason for self medication.

**Table 1.** Distribution of socio-demographic parameters on the basis of self-medication.

| Predictors | Do you give antibiotics to your child without prescription |  | Total<br>N =173<br>(%) |
| --- | --- | --- | --- |
|  | N=118<br>No (%) | N=55<br>Yes (%) |  |
| Relationship to child |  |  |  |
| Mother | 76 (43.9) | 43 (24.9) | **119 (68.8)** |
| Father | 42 (24.3) | 12 (6.9) | **54 (31.2)** |
| Age |  |  |  |
| <30 | 44 (25.4) | 16 (9.2) | **60 (34.7)** |
| 30-39 | 43 (24.9) | 27 (15.6) | **70 (40.5)** |
| ≥40 | 31 (17.9) | 12 (6.9) | **43 (24.9)** |
| Education |  |  |  |
| Secondary/ Higher secondary | 40 (23.1) | 05 (2.9) | **45 (26.0)** |
| Graduate | 53 (30.6) | 35 (20.2) | **88 (50.9)** |
| Post graduate | 25 (14.5) | 15 (8.7) | **40 (23.1)** |
| Family Income |  |  |  |
| ≤20000 | 10 (5.8) | 02 (1.2) | **12 (6.9)** |
| 20001-40000 | 60 (34.7) | 34 (19.7) | **94 (54.3)** |
| 40001-80000 | 38 (22.0) | 12 (6.9) | **50 (28.9)** |
| >80000 | 10 (5.8) | 07 (4.0) | **17 (9.8)** |
| Any Family member working in medical field? |  |  |  |
| No | 98 (56.6) | 36 (20.8) | **134 (77.5)** |
| Yes | 20 (11.6) | 19 (11.0) | **39 (22.5)** |
| Occupation |  |  |  |
| Housewife | 53 (30.6) | 31 (17.9) | **84 (48.6)** |
| Service | 42 (24.3) | 22 (12.7) | **64 (37.0)** |
| Business/ Skilled/Semi skilled worker | 23 (13.3) | 02 (1.2) | **25 (14.5)** |
| Ayushman Card |  |  |  |
| No | 117 (67.6) | 54 (31.2) | **171 (98.8)** |
| Yes | 01 (0.6) | 01 (0.6) | **02 (1.2)** |
| Child age |  |  |  |
| ≤3 | 24 (13.9) | 09 (5.2) | **33 (19.1)** |
| 4-6 | 38 (22.0) | 24 (13.9) | **62 (35.8)** |
| 7-9 | 35 (20.2) | 11 (6.4) | **46 (26.6)** |
| >9 | 21 (12.1) | 11 (6.4) | **32 (18.5)** |

**Table 2.** Behavior of parents towards self mediation in children.

| Predictors | Do you normally stop giving antibiotics when your child start feeling better? (N=173) |  | Do you keep antibiotic stocks at home for later use? (N=173) |  |
| --- | --- | --- | --- | --- |
|  | OR (95% CI) | p-value | OR (95% CI) | p-value |
| Relationship with child |  |  |  |  |
| Mother | Reference group |  | Reference group |  |
| Father | 0.945 (0.276 – 3.231) | 0.928 | 0.905 (0.237 – 3.461) | 0.884 |
| Age |  |  |  |  |
| <30 | Reference group |  | Reference group |  |
| 30-39 | 1.793 (0.687 – 4.680) | 0.233 | 0.912 (0.336 – 2.477) | 0.856 |
| ≥40 | 3.223 (0.878 – 11.828) | 0.078 | 0.842 (0.224 – 3.165) | 0.799 |
| Education |  |  |  |  |
| Secondary/ Higher secondary | Reference group |  | Reference group |  |
| Graduate | 0.159 (0.055 – 0.456) | 0.001 | 10.997 (3.749 – 32.254) | 0.000 |
| Post graduate | 0.863 (0.176 – 4.242) | 0.856 | 12.550 (3.192 – 49.338) | 0.000 |
| Family Income |  |  |  |  |
| ≤20000 | Reference group |  | Reference group |  |
| 20001-40000 | 1.211 (0.290 – 5.060) | 0.793 | 1.543 (0.292 – 8.170) | 0.610 |
| 40001-80000 | 0.735 (0.157 – 3.455) | 0.697 | 0.873 (0.153 – 4.990) | 0.879 |
| >80000 | 1.132 (0.177 – 7.231) | 0.896 | 0.692 (0.090 – 5.304) | 0.723 |
| Any Family member working in medical field? |  |  |  |  |
| No | Reference group |  | Reference group |  |
| Yes | 1.041 (0.392 – 2.765) | 0.936 | 4.780 (1.613 – 14.162) | 0.005 |
| Occupation |  |  |  |  |
| Housewife | Reference group |  | Reference group |  |
| Service | 1.400 (0.728 – 2.694) | 0.314 | 1.426 (0.461 – 4.412) | 0.538 |
| Business/Skilled/ Semi skilled worker | 0.280 (0.075 – 1.041) | 0.057 | 0.095 (0.014 – 0.632) | 0.015 |
| Ayushman Card |  |  |  |  |
| No | Reference group |  | Reference group |  |
| Yes | 0.751 (0.030 – 18.664) | 0.862 | 3.033 (0.003 – 3245.152) | 0.755 |
| Child's Age |  |  |  |  |
| <3 | Reference group |  | Reference group |  |
| 4-6 | 0.860 (0.297 – 2.492) | 0.782 | 0.867 (0.286 – 2.629) | 0.801 |
| 7-9 | 1.034 (0.308 – 3.467) | 0.957 | 0.447 (0.127 – 1.573) | 0.210 |
| >9 | 0.726 (0.173 – 3.051) | 0.662 | 0.484 (0.109 – 2.150) | 0.340 |

OR—Odds ratio; CI—Confidence interval

**Table 3.** Behavior of parents on the basis of self medication and Age.

| Variables | Do you give antibiotics to your child without prescription |  | Age of parents |  |  | Total (N=173) (%) |
| --- | --- | --- | --- | --- | --- | --- |
| | No<br>N (%) | Yes<br>N (%) | <30<br>N (%) | 30 – 39<br>N (%) | $\geq 40$<br>N (%) | |
| **Could identify antibiotic correctly** |  |  |  |  |  |  |
| No | 100 (57.8) | 04 (2.3) | 37 (21.4) | 38 (22.0) | 29 (16.8) | **104 (60.1)** |
| Yes | 18 (10.4) | 51 (29.5) | 23 (13.3) | 32 (18.5) | 14 (8.1) | **69 (39.9)** |
| **How long do you wait before starting antibiotic when your children get sick<sup>#</sup>** |  |  |  |  |  |  |
| Immediate | 12 (6.9) | 04 (2.3) | 06 (3.5) | 08 (4.6) | 02 (1.2) | **16 (9.2)** |
| 1-2 days | 98 (56.6) | 49 (28.3) | 50 (28.9) | 58 (33.5) | 39 (22.5) | **147 (85.0)** |
| 3-5 days | 08 (4.6) | 02 (1.2) | 04 (2.3) | 04 (2.3) | 02 (1.2) | **10 (5.8)** |
| **Reasons for self-medication<sup>#</sup>** |  |  |  |  |  |  |
| Previous experience with similar symptoms | 01 (0.6) | 37 (21.4) | 11 (6.4) | 17 (9.8) | 10 (5.8) | **38 (22.0)** |
| Minor Illness | 01 (0.6) | 43 (24.9) | 14 (8.1) | 18 (10.4) | 12 (6.9) | **44 (25.4)** |
| Long waiting time at clinics | 00 (00) | 02 (1.2) | 00 (00) | 01 (0.6) | 01 (0.6) | **02 (1.2)** |
| High cost of medical consultation | 00 (00) | 01 (0.6) | 01 (0.6) | 00 (00) | 00 (00) | **01 (0.6)** |
| Knowledge as a healthcare professional | 00 (00) | 09 (5.2) | 02 (1.2) | 06 (3.5) | 01 (0.6) | **09 (5.2)** |

<sup>#</sup>Multiple answer choice

## Discussion

The decision-making process of parents regarding the self-medication of antibiotics among children presents a critical and complex aspect of healthcare management within family dynamics. The present study included 173 parents residing in the study population area, of which, mothers were more involved in self-medicating their children with antibiotics which was similar to another study.^6^ Higher prevalence of self-medication was found among parents between 30-39 years age group whereas another study by Tibdewal and Gupta revealed prevalence of self-medication was more among parents of 21-34 years.^7^ Middle income families were seen to practice self medication more as also seen in other study.^1^ In our study, prevalence of self medication was significantly high among those who do not have Ayushman card (31.2%). Parental self-medication was found to be significantly more in children aged 4-6 years and above which was consistent with other study.^8^

Our result showed that mothers, parents ≥40 years age, having Secondary/ Higher secondary education normally stop giving antibiotics when their child starts feeling better whereas, Sharif et al.^9^ in another study showed that almost 50% participants stopped administration of antibiotic on completion of the course and only 9% stopped after a few days of recovery. Parents of <30 years, doing service, having postgraduate degree, having Ayushman card, and having family member working in medical field were seen keeping antibiotic stock at home for later use. A systematic review^4^ revealed that around 40%, ranging from 12% to 80.5% parents believed that leftover antibiotics in the past can be used to treat their children. This was statistically associated with higher risk of antibiotic self medication with an odds ratio of 3.01.

In our study, among those participants who could identify antibiotics correctly, only 29.5% parents self medicate their children, in contrast to a Chinese study by Lin L. et al. which showed parents with a medium or high ability to recognize antibiotics tended to self-medicate their children.^10^ Most common reason for self medication in this study was minor illness. Similar finding was seen in other studies,^11 12^ whereas a study by Tareq L. Mukattash et al. revealed previous experience with antibiotic efficacy and their perception that symptoms do not require a physician consultation was the main reason.^13^

## Conclusion

About one third of the participants practiced self medication in this study. The present study emphasizes on the need for comprehensive educational campaigns aimed at enhancing parental awareness regarding the risks associated with antibiotic misuse and the importance of seeking professional medical advice. Furthermore, fostering effective communication channels between healthcare providers and parents can facilitate informed decision-making and promote responsible antibiotic usage. By addressing misconceptions, empowering parents with knowledge and promoting prudent antibiotic stewardship practices, we can collectively strive towards safeguarding the health and well-being of our children and future generations.

## Data availability statement

The data that support the findings of this study are available from the corresponding author upon reasonable request.

## Financial support and sponsorship

Nil.

## Conflicts of interest

There are no conflicts of interest.

## Acknowledgements

None.

## Notes

### Competing Interest Statement

The authors have declared no competing interest.

### Funding Statement

This study did not receive any funding

### Author Declarations

The study started after receiving approval (Letter No./807/IEC/IGIMS/2022) from the Institutional Human Ethics Committee and the Dean of Research at the Indira Gandhi Institute of Medical Sciences, Patna.

